# Association between angiotensin-converting enzyme inhibitors and angiotensin II receptor blockers use and the risk of infection and clinical outcome of COVID-19: a comprehensive systematic review and meta-analysis

**DOI:** 10.1101/2020.07.02.20144717

**Authors:** Guangbo Qu, Liqin Shu, Evelyn J. Song, Dhiran Verghese, John Patrick Uy, Ce Cheng, Qin Zhou, Hongru Yang, Zhichun Guo, Mengshi Chen, Chenyu Sun

**Affiliations:** Department of Epidemiology and Health Statistics, School of Public Health, Anhui Medical University, No. 81 Meishan Road, Hefei 230032, Anhui, China; Department of Child Health care, Maternal and Child Health Care Hospital of Anhui Province (Affiliated Maternal and Child Health Care Hospital of Anhui Medical University), Hefei 230001, Anhui, China; Department of Medicine, School of Medicine, Johns Hopkins University, Baltimore, MD, USA; AMITA Health Saint Joseph Hospital Chicago, 2900 N. Lake Shore Drive, Chicago, 60657, Illinois, USA; Department of Internal Medicine, Cape Fear Valley Medical Center, Fayetteville 28304, NC, USA; Mayo Clinic, Rochester, MN 55905, USA; Massachusetts College of Pharmacy and Health Science, 179 Longwood Ave, Boston, 02115, MA, USA; Hunan Provincial Key Laboratory of Clinical Epidemiology, Xiangya School of Public Health, Central South University

**Keywords:** COVID-19, Angiotensin-converting enzyme inhibitor, Angiotensin-receptor blockers, risk, systematic review, meta-analysis

## Abstract

**Background:** The effect of using Angiotensin-converting enzyme inhibitors (ACEIs) and Angiotensin-receptor blockers (ARBs) on the risk of coronavirus disease 2019 (COVID-19) is a topic of recent debate. Although studies have examined the potential association between them, the results remain controversial. This study aims to determine the true effect of ACEI/ARBs use on the risk of infection and clinical outcome of COVID-19.

**Methods:** Five electronic databases (PubMed, Web of science, Cochrane library, China National Knowledge Infrastructure database, medRxiv preprint server) were retrieved to find eligible studies. Meta-analysis was performed to examine the association between ACEI/ARBs use and the risk of infection and clinical outcome of COVID-19.

**Results:** 22 articles containing 157,328 patients were included. Use of ACEI/ARBs was not associated with increased risk of infection (Adjusted OR: 0.96, 95% CI: 0.91-1.01, I^2^=5.8%) or increased severity (Adjusted OR: 0.90, 95% CI: 0.77-1.05, I^2^=27.6%) of COVID-19. The use of ACEI/ARBs was associated with lower risk of death from COVID-19 (Adjusted OR: 0.66, 95% CI: 0.44-0.99, I^2^=57.9%). Similar results of reduced risk of death were also found for ACEI/ARB use in COVID-19 patients with hypertension (Adjusted OR: 0.36, 95% CI: 0.17-0.77, I^2^=0).

**Conclusion:** This study provides evidence that ACEI/ARBs use for COVID-19 patients does not lead to harmful outcomes and may even provide a beneficial role and decrease mortality from COVID-19. Clinicians should not discontinue ACEI/ARBs for patients diagnosed with COVID-19 if they are already on these agents.

## 1. Introduction

In December 2019, the first case of novel coronavirus disease (COVID-19) caused by the severe acute respiratory syndrome coronavirus 2 (SARS-CoV-2) was reported in Wuhan, China [1, 2]. Due to the rapidly increasing number of cases worldwide, on March 11, 2020, the World Health Organization (WHO) declared the COVID-19 outbreak a global pandemic As of June 3, 2020, there were 6, 287, 771 confirmed cases and 379, 941 deaths globally per WHO [3]. COVID-19 has greatly impacted both professional and personal lives of everyone worldwide.

The epidemiological and clinical characteristics of COVID-19 have been well described in previous studies [4, 5]. Hypertension, diabetes and cardiovascular diseases (CVD) including congestive heart failure (CHF) and myocardial infarction are common comorbidities reported in patients with COVID-19 and have been associated with a poor prognosis [5, 6]. Angiotensin-converting enzyme inhibitors (ACEIs) and Angiotensin-receptor blockers (ARBs) are first line anti-hypertensive medications and are included in the guideline-directed therapy for diabetic nephropathy, CHF and myocardial infarction. Angiotensin-converting enzyme 2 (ACE2), a membrane-bound aminopeptidase, is widely expressed in certain tissues of human body including lung, intestine, heart, and kidneys, and it has been shown that the use of ACEIs and ARBs can increase the expression of ACE2 [7, 8]. Furthermore, ACE2 acts as a functional receptor and SARS-CoV-2 utilizes the ACE2 for attachment of its spike protein, in the process of its entrance into cells [9,10]. These findings led some investigators to hypothesize that the use of ACEIs or ARBs may increase the risk of infection by SARS-CoV-2 and the severity of COVID-19 [11-13].

Several clinical studies have been conducted to test this hypothesis. However, a consensus has not been reached regarding how the use of ACEIs or ARBs affects the outcome of patients with COVID-19. The goal of this meta-analysis is to clarify the effects of ACEIs and/or ARBs on the infection risk, severity, and mortality of COVID-19, hoping to shed more light in the prevention and treatment of the current ongoing pandemic.

## 2. Methods

This study was conducted and reported according to the Preferred Reporting Items for Systematic Reviews and Meta-Analyses (PRISMA) statement [14].

### 2.1 Literature search and data source

We searched five electronic databases (PubMed, Web of science, Cochrane library, China National Knowledge Infrastructure database, medRxiv preprint server) to collect relevant studies published until May 20, 2020. The search strategy was established and performed by two authors. The search terms used included the following: (angiotensin-converting-enzyme inhibitor OR ACEI OR angiotensin-receptor blockers OR ARB) AND (coronavirus OR COVID-19 OR SARS-CoV-2). There was no language limitation on the search. All references of included studies were evaluated for additional studies to include as many eligible studies as possible. For retrieved records, we used reference management software (NoteExpress, college version) to save and filter.

### 2.2 Study selection

First, we reviewed the title and abstract of retrieved studies and excluded irrelevant ones. Then, we read through the full-text of remaining studies and included eligible studies based on our inclusion and exclusion criteria. Our inclusion criteria are as follows: (1) participants were patients diagnosed with COVID-19; (2) patients were reported as having ACEI and/or ARB exposure and non-ACEI and/or ARB exposure (ACEI use versus non-ACEI use; ARB use versus non-ARB use; ACEI/ARB use versus non-ACEIARB use); (3) at least one of following outcomes of COVID-19 was reported: positive rate/infection rate, hospital admission rate, severity, mortality in groups of ACEI and/or ARB exposure and non-ACEI and/or ARB exposure or the association between ACEI and/or ARB exposure and the risk of infection, hospital admission, severity, and death due to COVID-19; (4) the study design was case-control or cohort. Exclusion criteria are as follows: (1) the studies were reviews, case-reports, or animal experiments; (2) studies did not meet the inclusion criteria; (4) The data of interest was not reported; (4) no clear definition or diagnose methods of COVID-19. Two authors (Guangbo Qu and Liqin Shu) conducted the selections independently, and any disagreement was solved through discussion.

### 2.3 Data extraction and quality assessment

Following information was extracted using a standard chart form: first author’s name, number of patients, number of patients in ACEI/ARB exposure (only ACEI exposure, only ARB exposure, either ACEI or ARB exposure) and non-exposure groups, patients’ characteristics, infection risk, hospital admission, severity, and mortality of patients in each group, odds ratios (ORs) or risk ratios (RRs) or hazard ratio (HRs) of the infection, hospital admission, severity, and mortality of COVID-19.

Considering that the included studies were observational studies, the Newcastle-Ottawa Scale (NOS) was used to assess the quality from three aspects: (1) selection of participants; (2) comparability of groups; (3) assessment of exposure and outcome [15]. NOS contained eight items with scores ranging from 0 to 9 stars. The quality of studies was divided into three categories based on the scores: low quality is 0-3 stars, moderate quality is 4-5 stars, and high quality is 6 stars or above. Two authors (Guangbo Qu and Liqin Shu) assessed quality independently, and disagreement was resolved by consensus.

## 3. Statistical analysis

Meta-analyses were conducted to assess for associations. Crude and adjusted odds ratios (Crude OR and Adjusted OR), with corresponding 95% confidence intervals, were calculated using the extracted binary data or effect sizes that was reported in all studies. Additionally, if available, Crude OR and/or Adjusted OR were also calculated to explore the association between ACEI/ARB use and outcomes of COVID-19 among patients with hypertension. The statistical heterogeneity between studies was identified using the Cochrane’s Q test and I-square test. Heterogeneity was present if P value of Q test was less than 0.1 or the value of I^2^ was more than 50% [16]. The model for meta-analyses was chosen based on the level of heterogeneity between studies; if obvious heterogeneity was present, then random-effects model was used, otherwise, the fix-effects model was used [17]. Subgroup analysis was conducted based on the characteristics of included studies. Publication bias was identified via Begg’s and Egger’s tests. If P value of Begg’s and Egger’s tests were more than 0.05, then publication bias is present. Sensitivity analysis was performed by omitting studies one by one to assess the stability of pooled results. All statistical analyses were performed using STATA software (version 14.0) and Review Manager (version 5.3).

## 4. Results

### 4.1 Characteristics of included studies

After initial search, 477 records were retrieved (PubMed: n=166, Web of science: n=228, Cochrane library: n=9, China National Knowledge Infrastructure database: n=27, medRxiv preprint server: n=41, Other sources: n=6). 27 records were excluded for duplication. After reading the titles and abstracts, 391 records were excluded due to irrelevance. After study selection according to the inclusion and exclusion criteria, 36 articles were excluded and one study was further excluded because it was retracted by the journal. Finally, 22 articles containing 157,328 patients were included in this meta-analysis [8, 10, 18-37]. The flow chart of the selection process is displayed in Figure 1.

**Figure 1.**
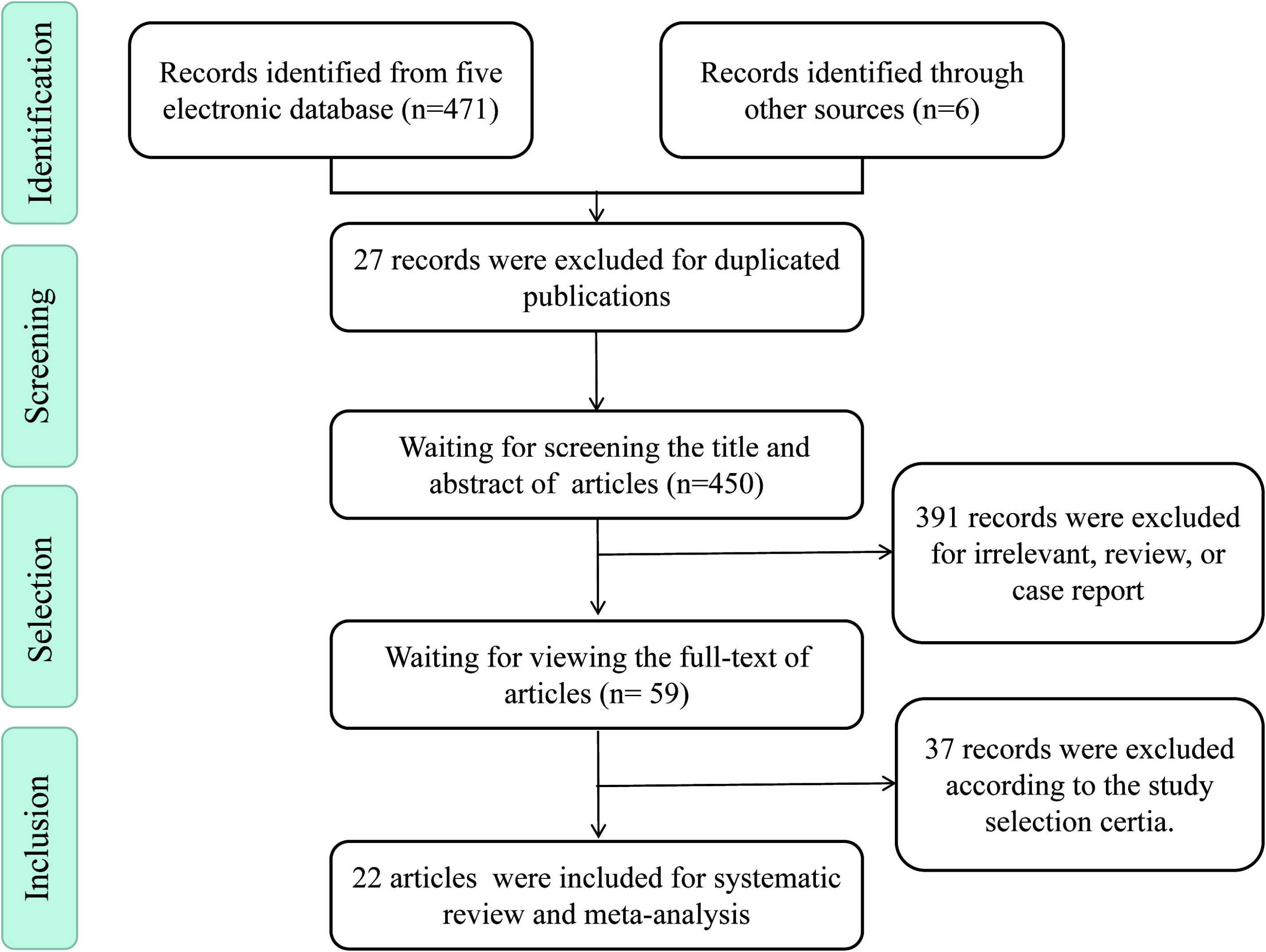
Flow diagram of study selection for this systematic review and meta-analysis.

The characteristics of included studies were shown in Table 1. Of included studies, 11 were conducted in China [8,16,20,21,22,24,25,27,33,35,36], six were from the United States [19,23,29,32,34,37], two were from Italy [10,30], one from Spain [26]. one from United 5 Kingdom [28], and one from Belgium [31]. All, but three, were cohort studies. The sample size of each study ranged from 42 to 37,031. Multiple comorbidities were reported and all patients in eight of the studies had hypertension [8,18,22,25,27,32,33,37]. Majority of the studies were high quality studies. Detailed quality assessment results are in Table S1.

**Table 1.**
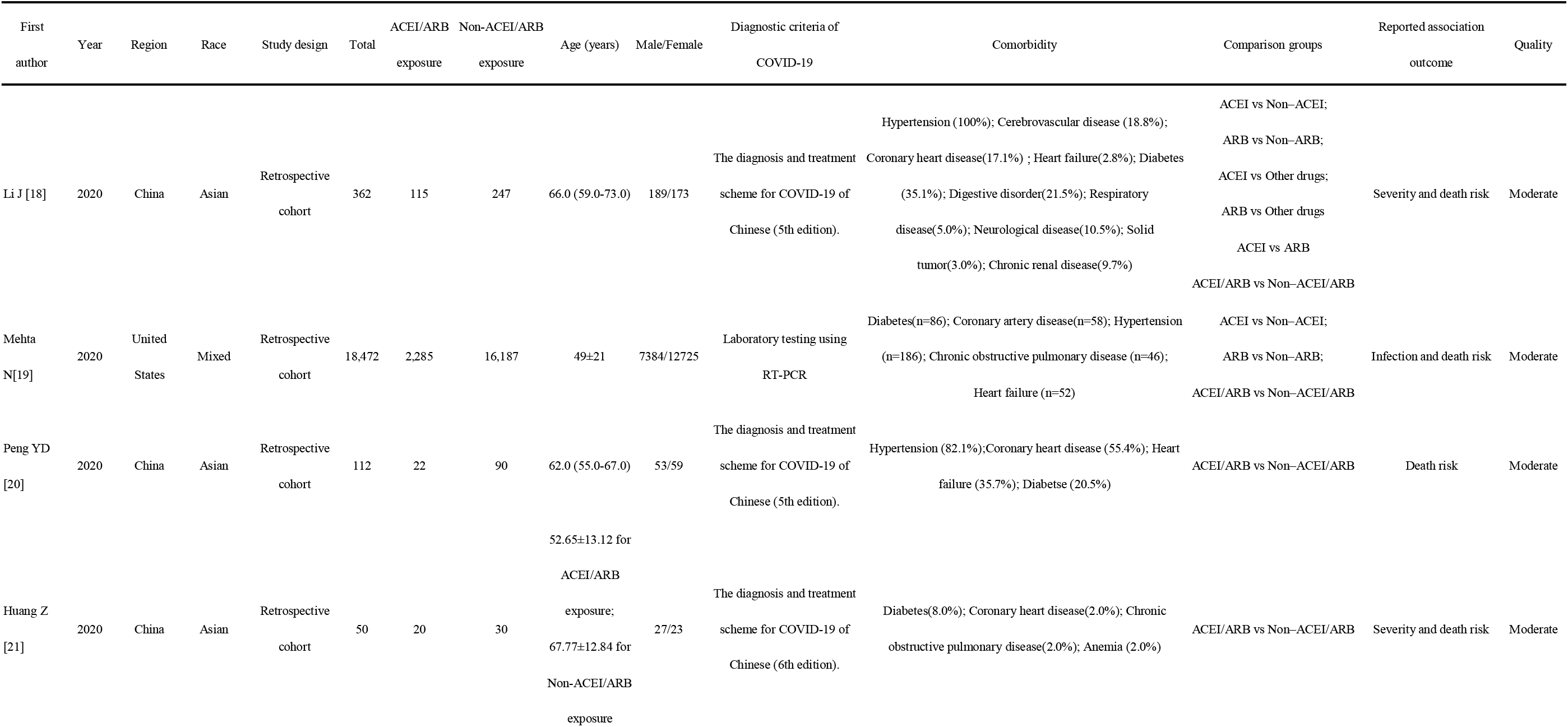
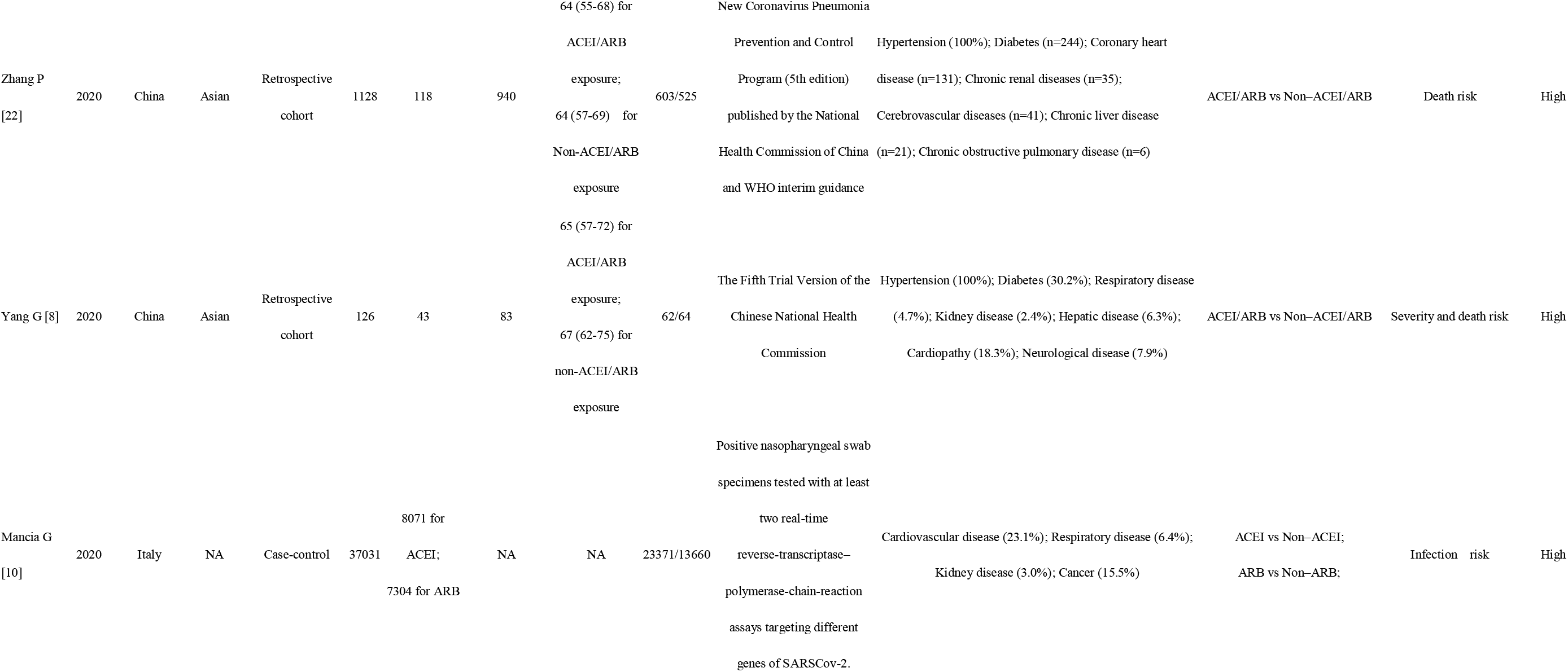
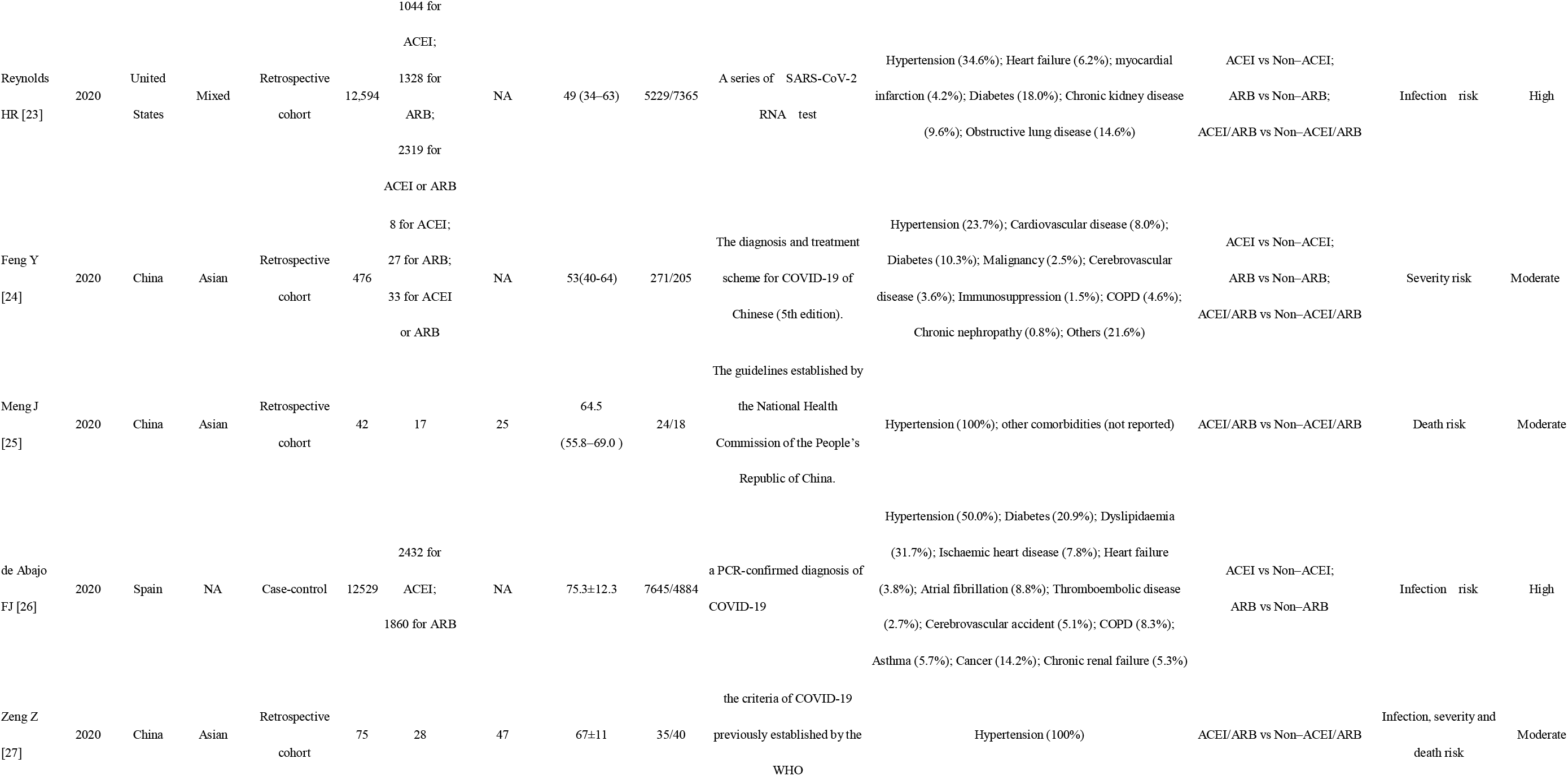
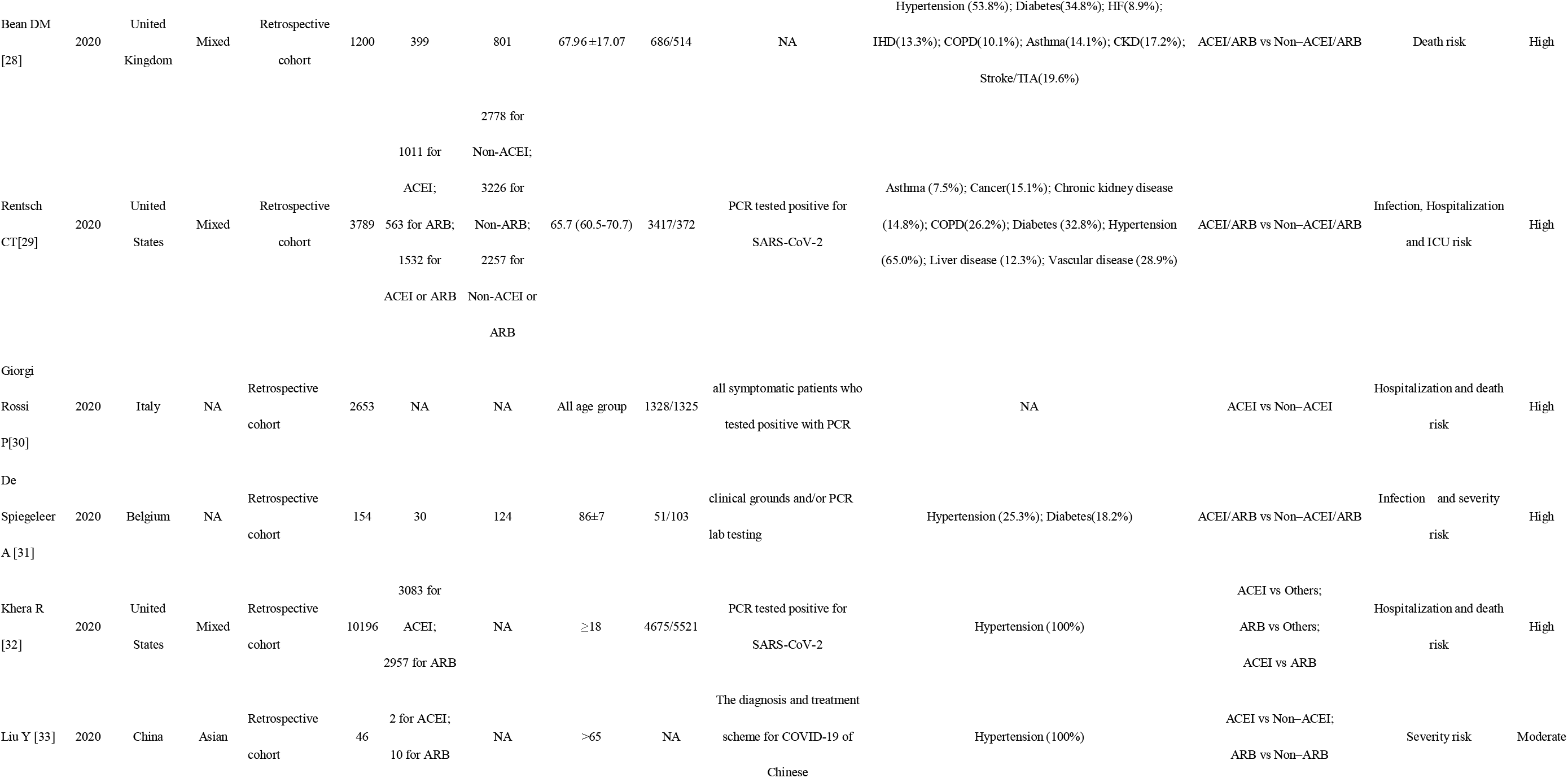
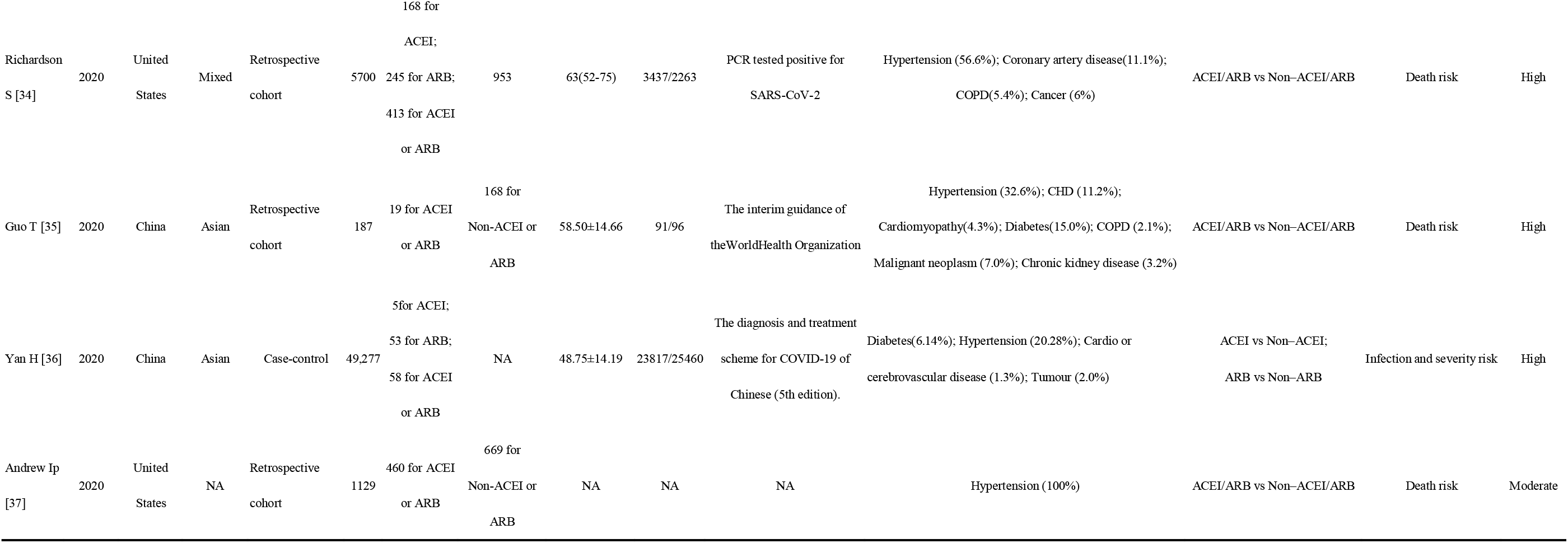
Characteristics of included studies.

### 4.2 ACEI/ARB use and COVID-19 infection

Pooled meta-analyses showed that, without adjusting for any confounders, ACEI, ARB, and ACEI/ARB use were not significantly associated with the risk of COVID-19 infection (Crude OR: 1.27, 95% CI: 0.95-1.69 for ACEI use; Crude OR: 1.07, 95% CI: 0.76-1.50 for ABR use; Crude OR: 1.10, 95% CI: 0.84-1.43 for ACEI/ARB use); however, obvious heterogeneity was present between studies (All I^2^ >50%) (Table 2, Figure 2). Similarly, after adjusting for confounders, ACEI, ARB, and ACEI/ARB use were not significantly associated with the risk of COVID-19 infection (Adjusted OR: 0.94, 95% CI: 0.87-1.01, I^2^=0 for ACEI use; Adjusted OR: 0.73, 95% CI: 0.49-1.09, I^2^=95.1% for ABR use; Adjusted OR: 0.96, 95% CI: 0.91-1.01, I^2^=5.8% for ACEI/ARB use). Furthermore, subgroup analysis based on different characteristics of studies showed no correlation between the use of ACEI and/or ARB and risk of COVID-19 infection.

**Table 2.**
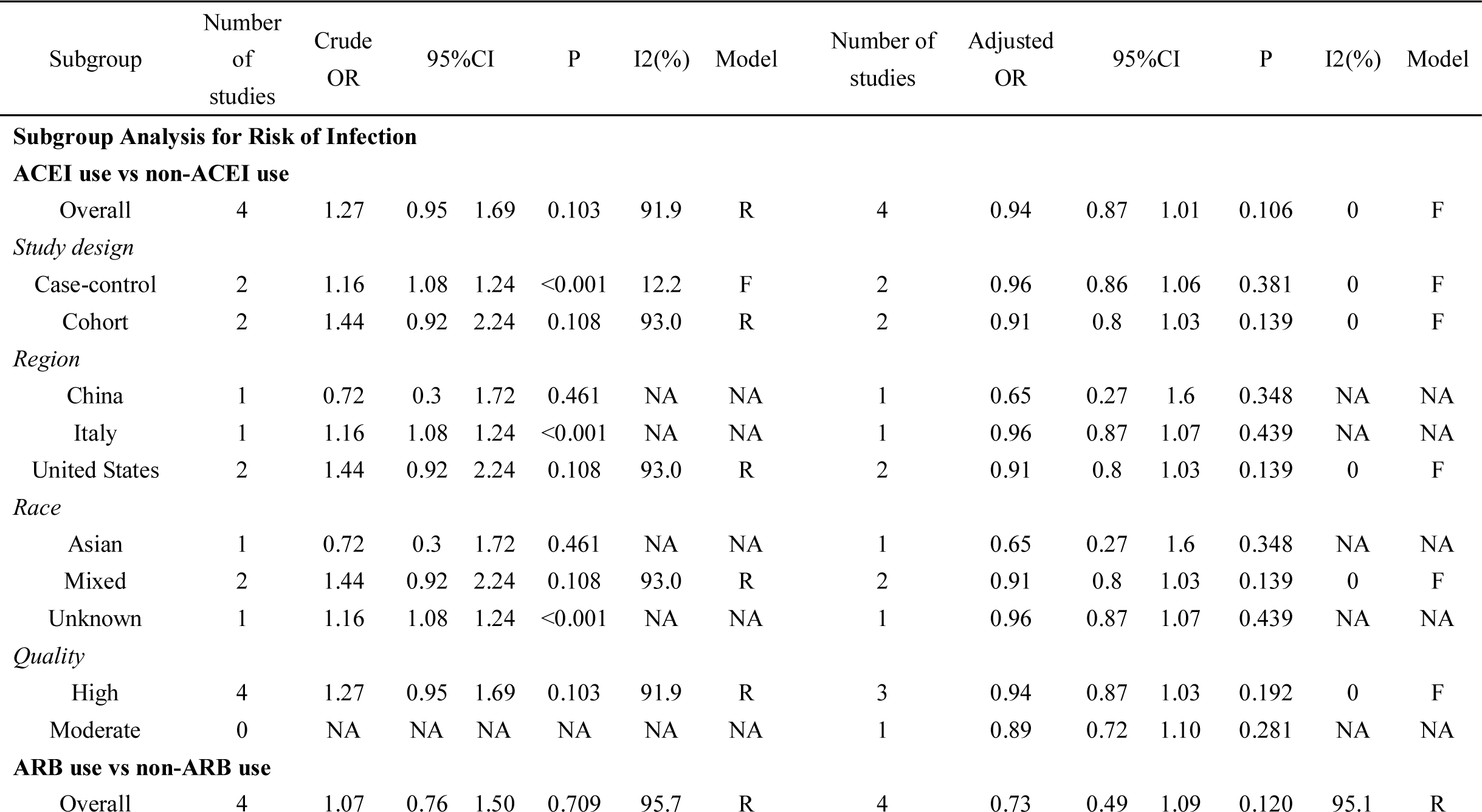
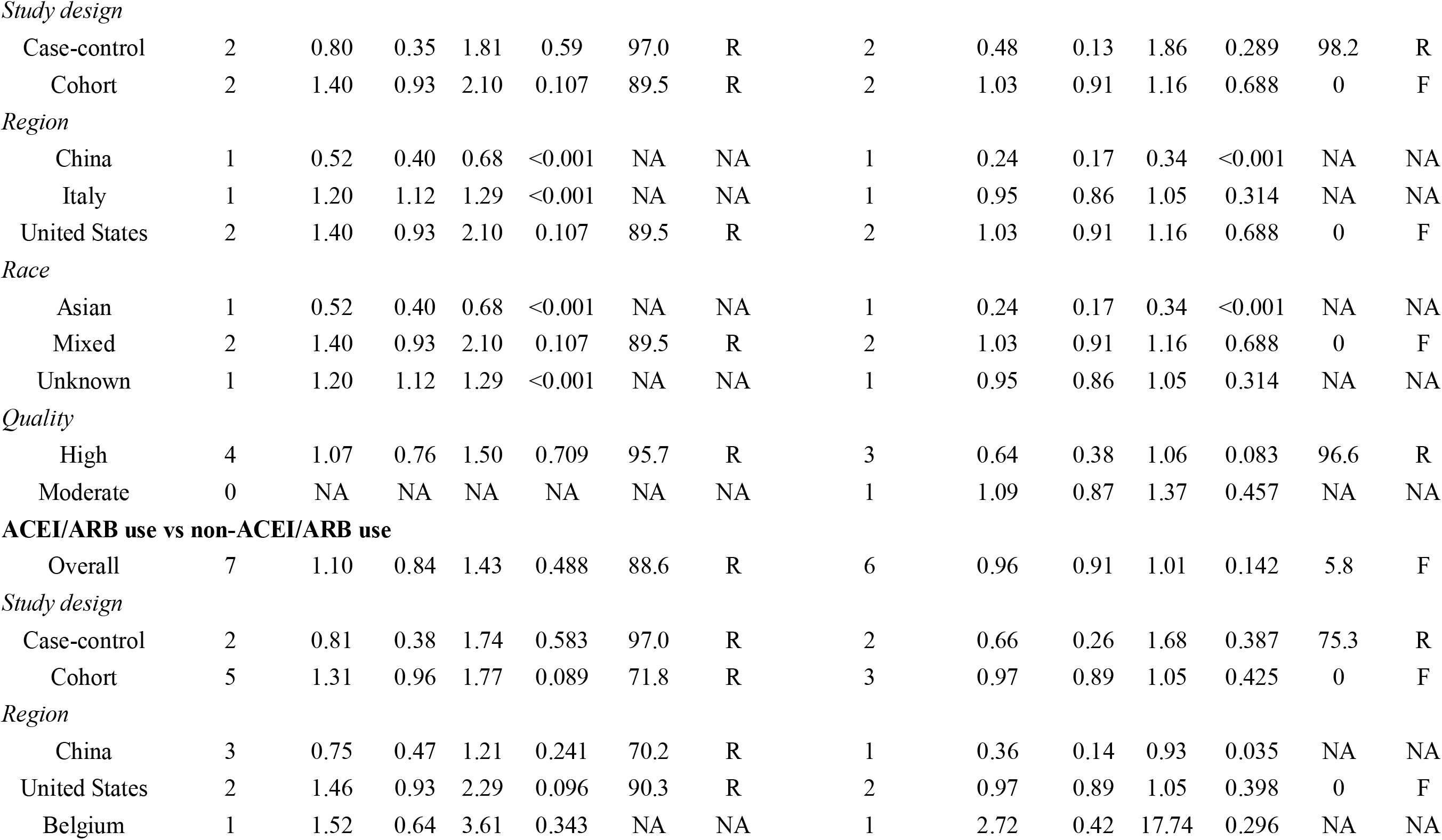
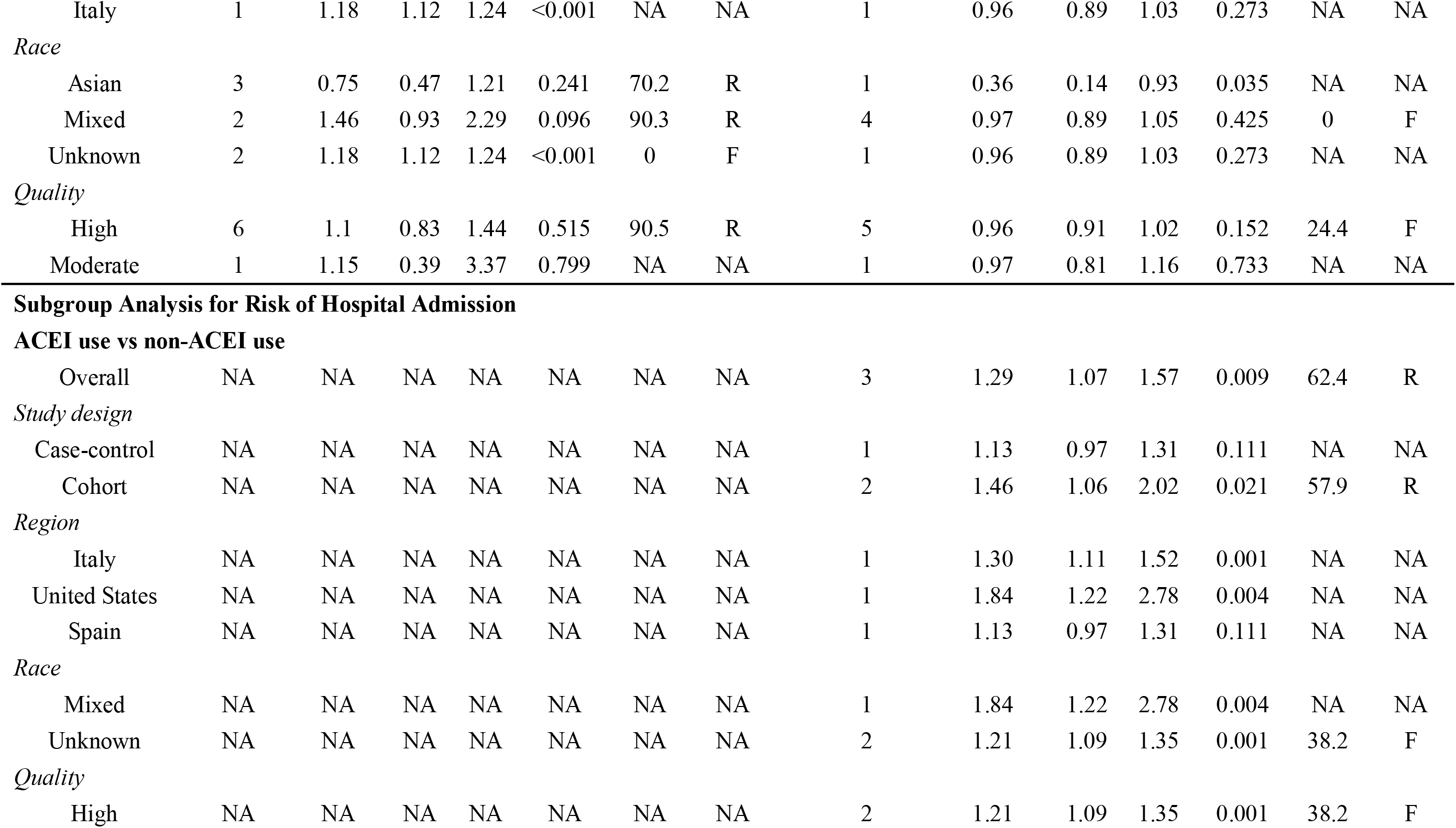
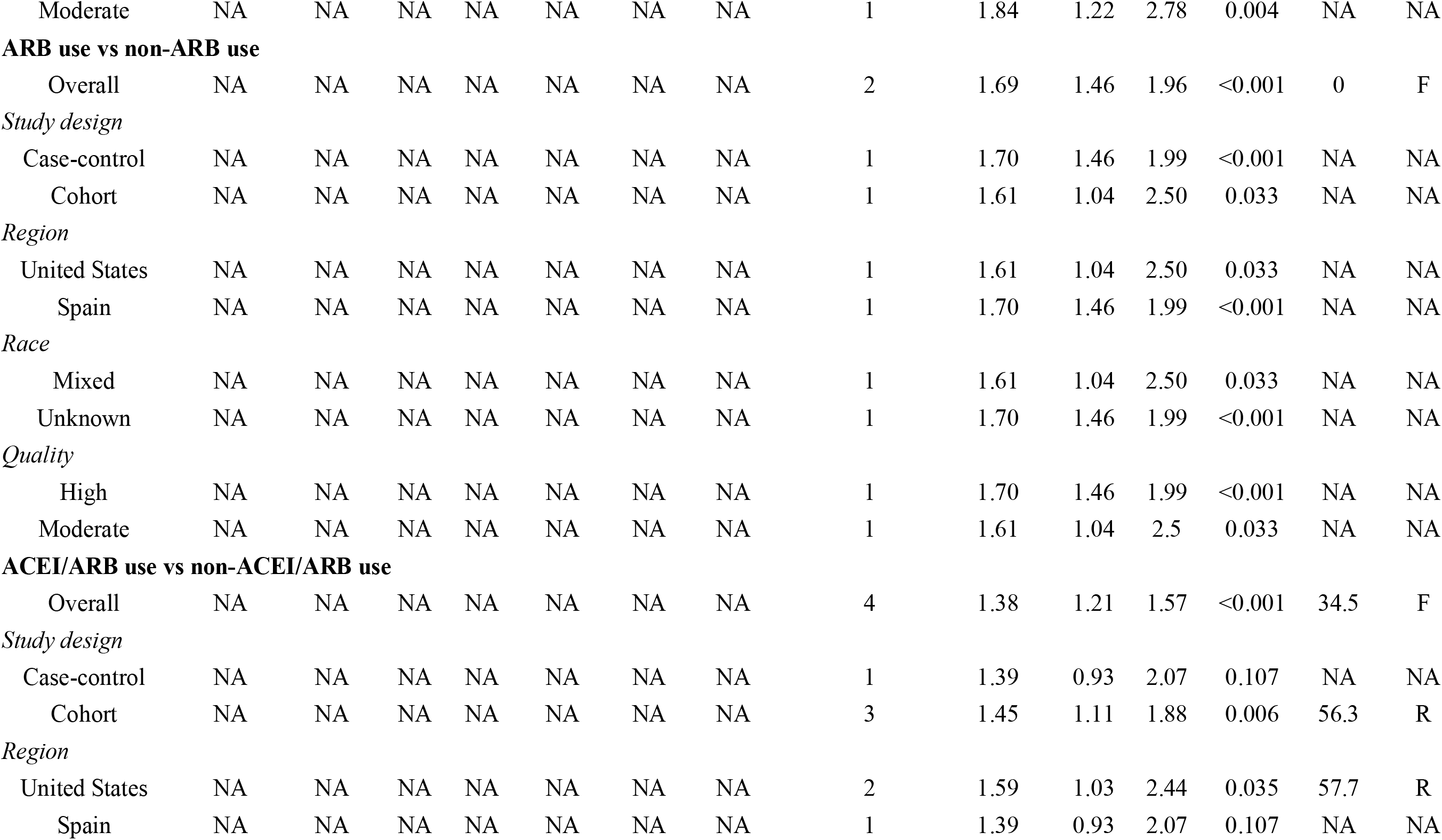
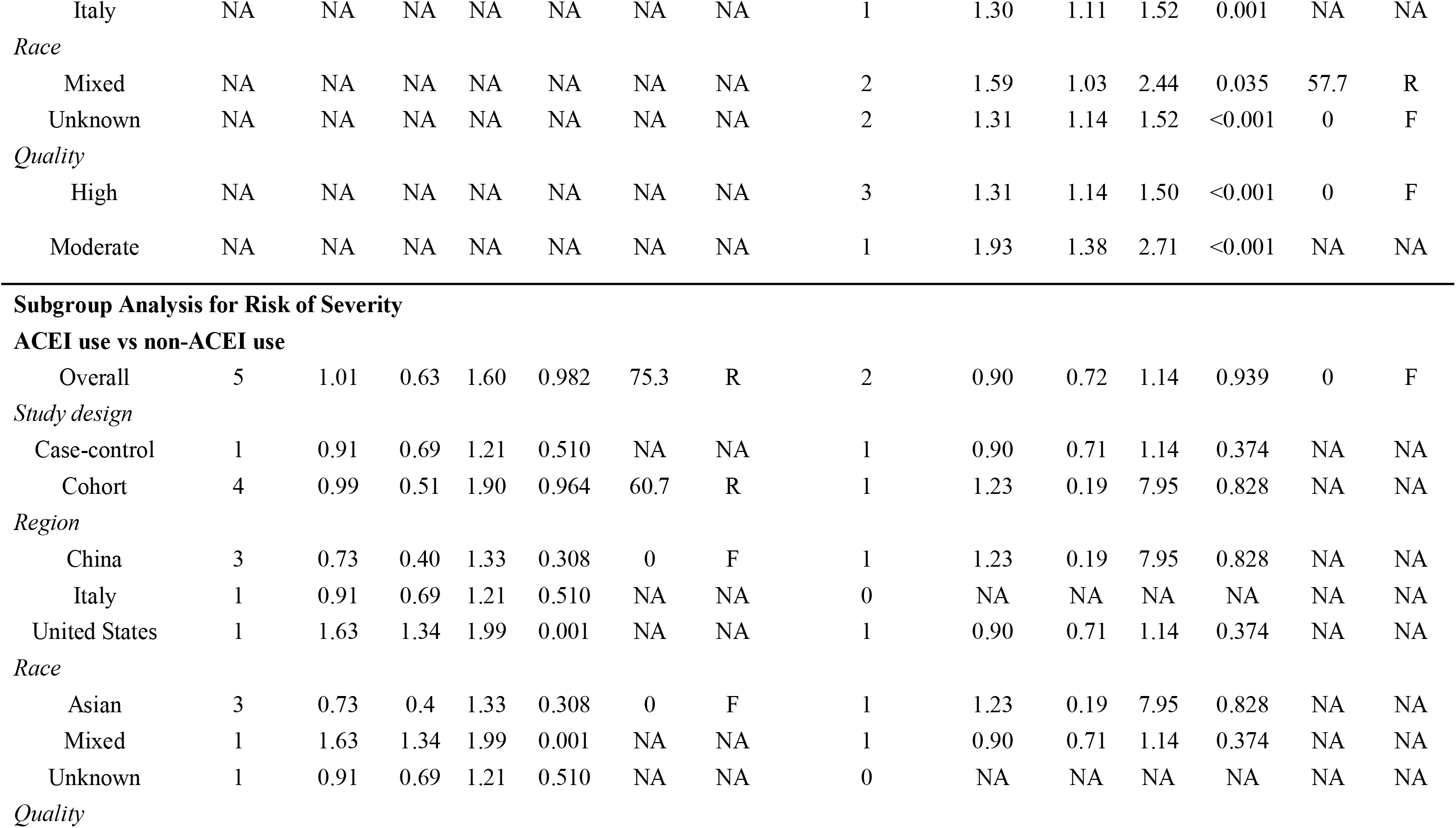
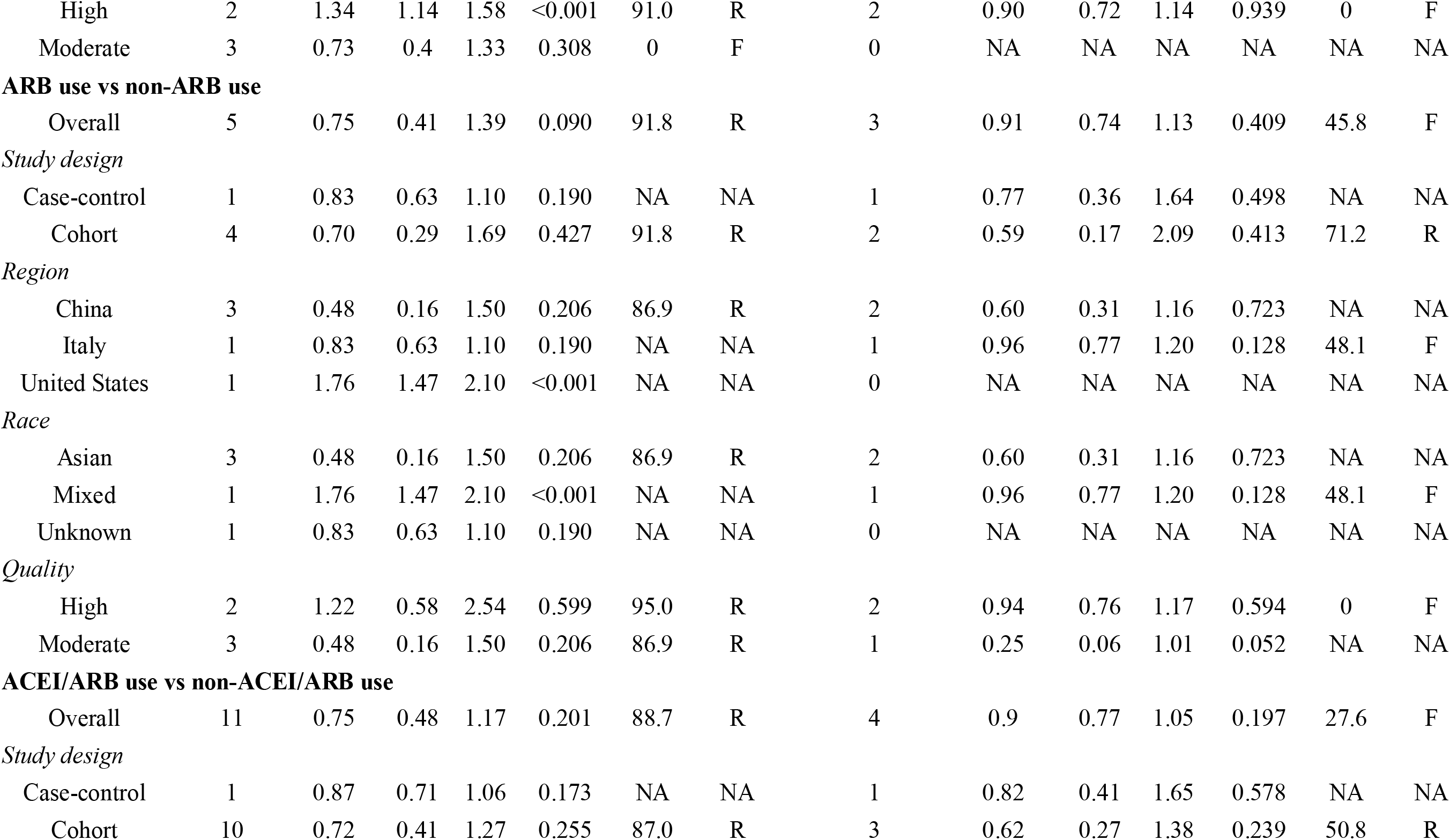
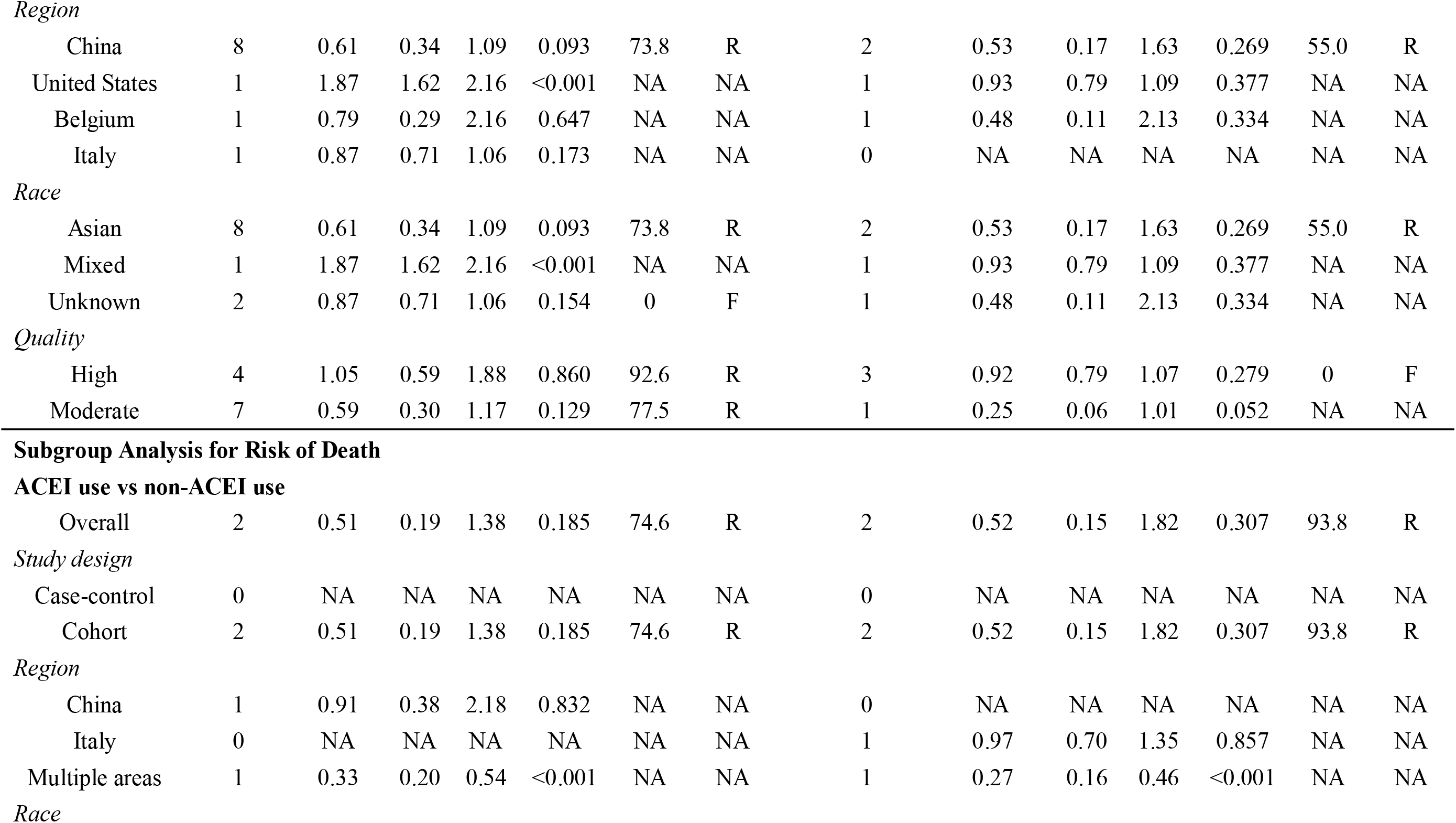
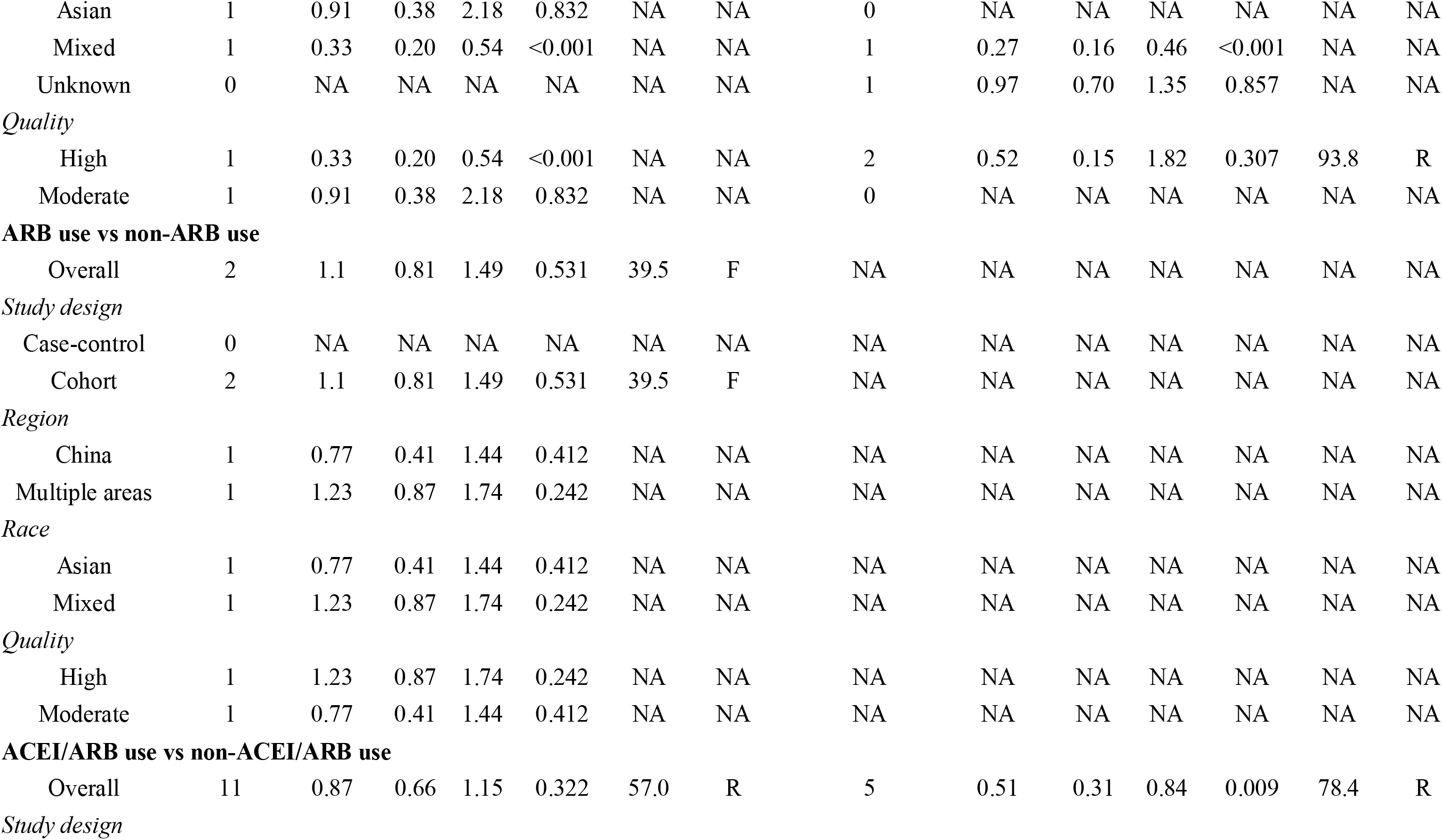
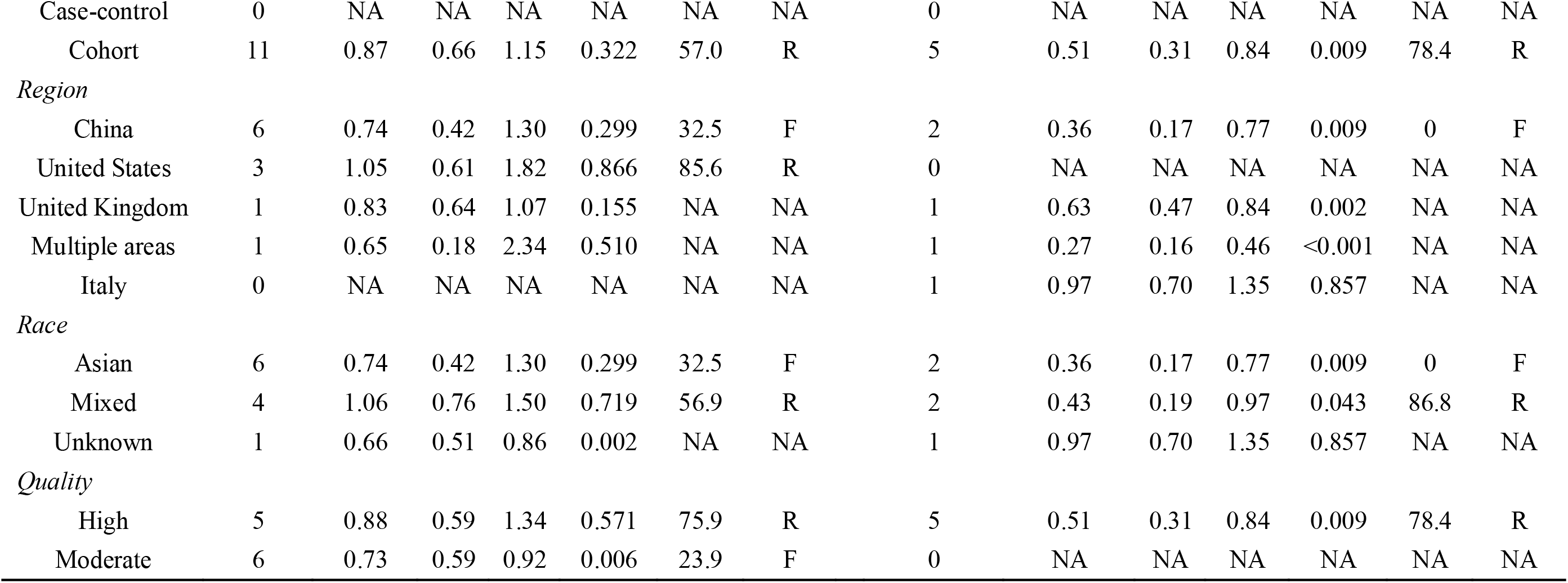
Meta-analysis on the association between ACEI/ARB use and risk of infection, hospital admission, severity, and death of COVID-19.

**Figure 2.**
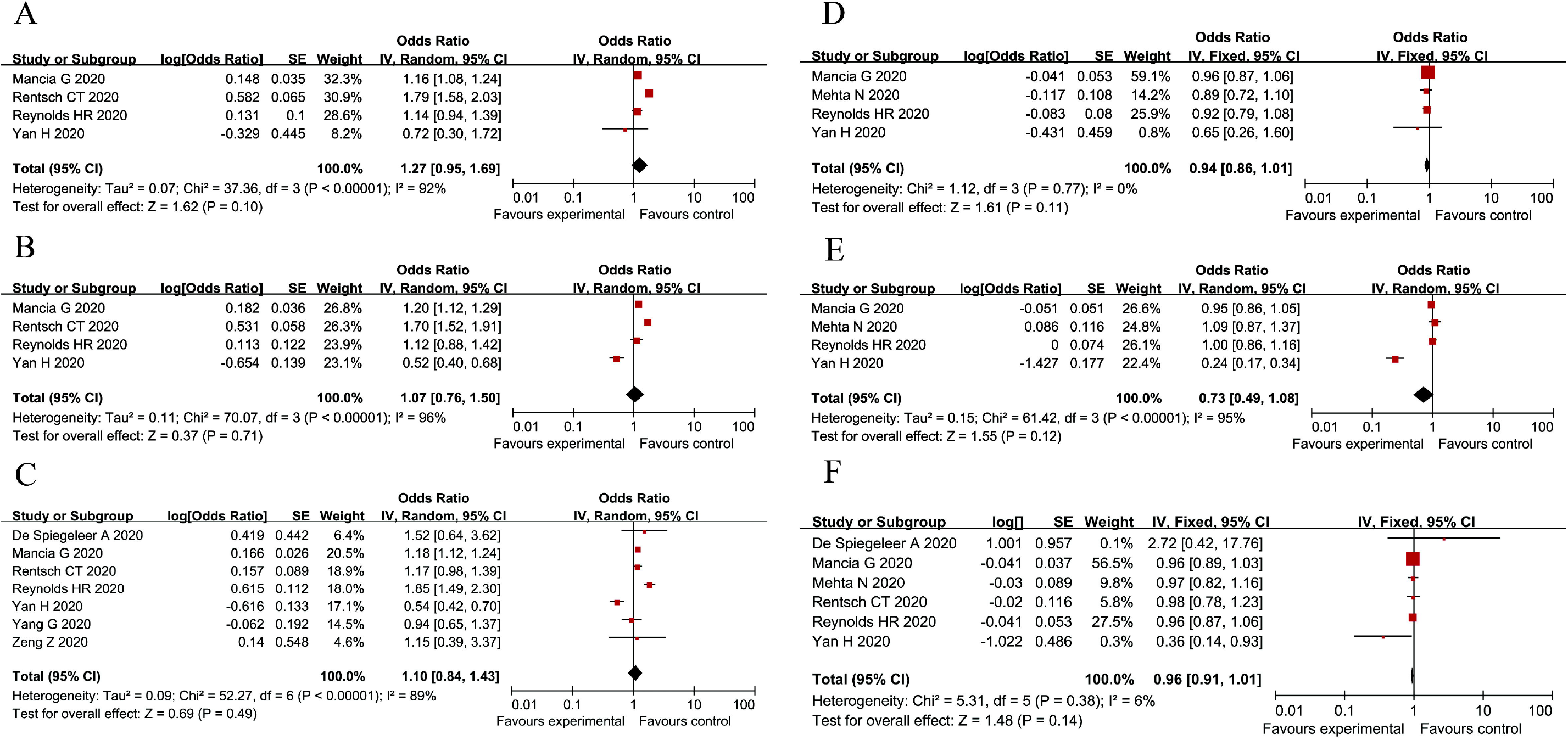
Pooled results of meta-analyses for the association between ACEI/ARB use and risk of COVID-19 infection. (A) Pooled crude OR for the comparison between ACEI use versus non-ACEI use; (B) Pooled crude OR for the comparison between ARB use versus non-ARB use; (C) Pooled crude OR for the comparison between ACEI/ARB use versus non-ACEI/ARB use; (D) Pooled adjusted OR for the comparison between ACEI use versus non-ACEI use; (E) Pooled adjusted OR for the comparison between ARB use versus non-ARB use; (F) Pooled adjusted OR for the comparison between ACEI/ARB use versus non-ACEI/ARB use.

### 4.3 ACEI/ARB use and hospital admission for COVID-19

Pooled meta-analyses showed that patients using ACEI, ARB, and ACEI/ARB had higher risk of hospital admission for COVID-19 compared to patients not using these medications (Adjusted OR: 1.29, 95% CI: 1.07-1.57, I^2^=62.4% for ACEI use; Adjusted OR: 1.69, 95% CI: 1.46-1.96, I^2^=0 for ABR use; Adjusted OR: 1.38, 95% CI: 1.21-1.57, I^2^=34.5% for ACEI/ARB use) (Table 2 and Figure 3).

**Figure 3.**
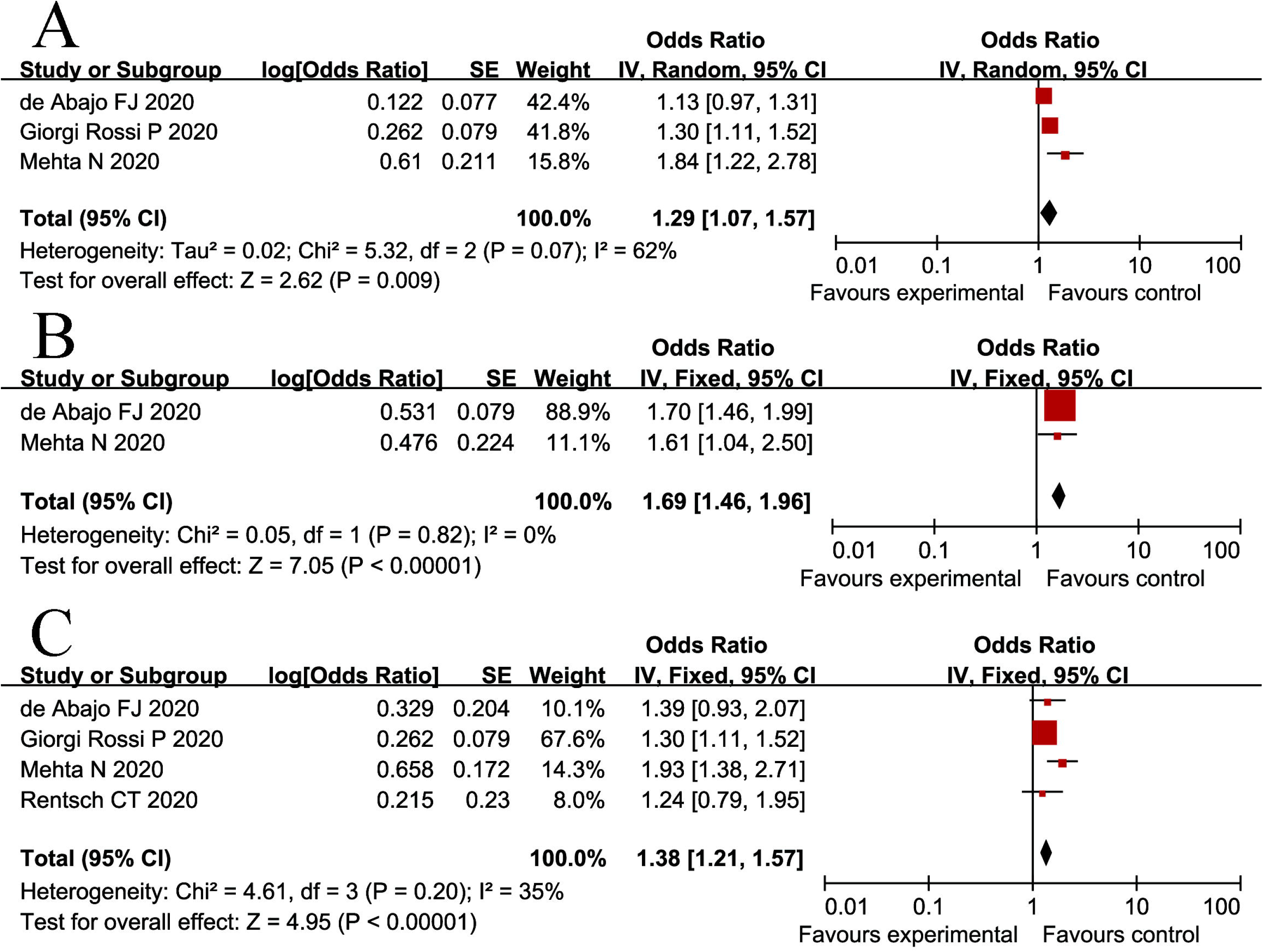
Pooled results of meta-analyses for the association between ACEI/ARB use and risk of hospital admission for COVID-19. (A) Pooled adjusted OR for the comparison between ACEI use versus non-ACEI use; (B) Pooled adjusted OR for the comparison between ARB use versus non-ARB use; (C) Pooled adjusted OR for the comparison between ACEI/ARB use versus non-ACEI/ARB use.

### 4.4 ACEI/ARB use and severity of COVID-19

Use of ACEI, ARB, and ACEI/ARB was not associated with severity of COVID-19 infection for both pooled crude and adjusted OR (Adjusted OR: 0.90, 95% CI: 0.72-1.14, I^2^=0 for ACEI use; Adjusted OR: 0.91, 95% CI: 0.74-1.13, I^2^=45.8% for ABR use; Adjusted OR: 0.90, 95% CI: 0.77-1.05, I^2^=27.6% for ACEI/ARB use). Additionally, subgroup analysis based on different characteristics (study design, region, race, study quality) also did not show any significant association (Table 2 and Figure 4). Furthermore, for patients with hypertension, ACEI, ARB, and ACEI/ARB use was not significantly associated with severity of COVID-19 infection (Table S2).

**Figure 4.**
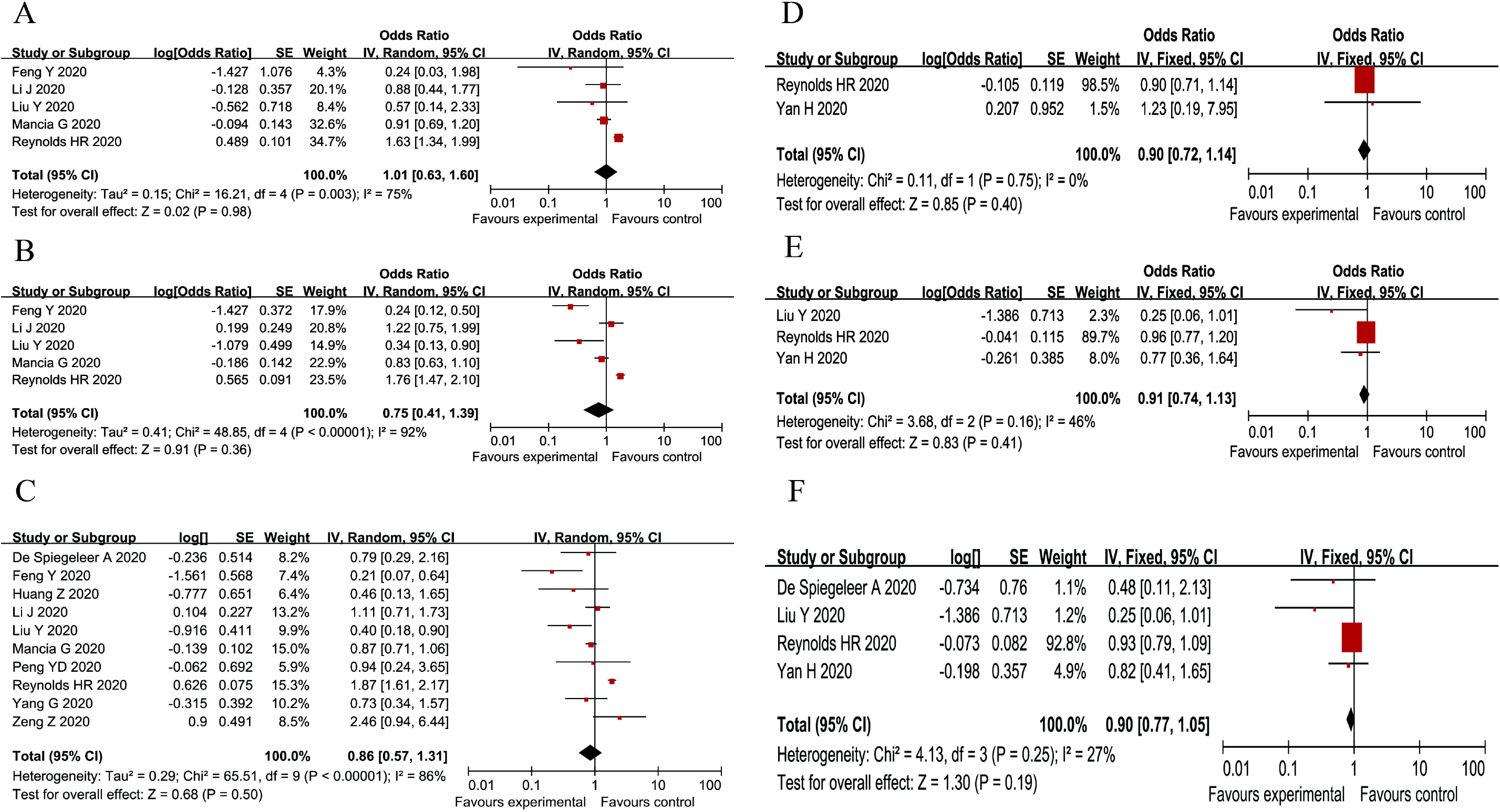
Pooled results of meta-analyses for the association between ACEI/ARB use and risk of severity of COVID-19 infection. (A) Pooled crude OR for the comparison between ACEI use versus non-ACEI use; (B) Pooled crude OR for the comparison between ARB use versus non-ARB use; (C) Pooled crude OR for the comparison between ACEI/ARB use versus non-ACEI/ARB use; (D) Pooled adjusted OR for the comparison between ACEI use versus non-ACEI use; (E) Pooled adjusted OR for the comparison between ARB use versus non-ARB use; (F) Pooled adjusted OR for the comparison between ACEI/ARB use versus non-ACEI/ARB use.

### 4.5 ACEI/ARB use and COVID-19 related mortality

Only one study reported the association between ACEI and ARB use and risk of death from COVID-19 without adjusting confounders separately (OR: 0.91, 95% CI: 0.38-2.17 for ACEI use; OR: 0.77, 95% CI: 0.41-1.43 for ABR use) [18]. Only one study reported no significant association between ACEI use and risk of death from COVID-19 (OR: 0.97, 95% CI: 0.69-1.34) [30]. The pooled crude OR showed no significant association between ACEI/ARB use and risk of death from COVID-19 (Crude OR: 0.88, 95% CI: 0.66-1.18, I^2^=60.9%). However, the pooled adjusted ORs showed that patients on ACEI/ARB had a lower risk of death from COVID-19 (Adjusted OR: 0.66, 95% CI: 0.44-0.99, I^2^=57.9%). No study reported the association between ARB use and the risk of death from COVID-19 (Table 2 and Figure 5). Furthermore, subgroup analysis on studies from China or studies of Asian population found that ACEI/ARB use was associated with a lower risk of death from COVID-19. For patients with hypertension, ACEI/ARB use also was associated with a lower risk of death from COVID-19 (Adjusted OR: 0.36, 95% CI: 0.17-0.77, I^2^=0) (Table S3).

**Figure 5.**
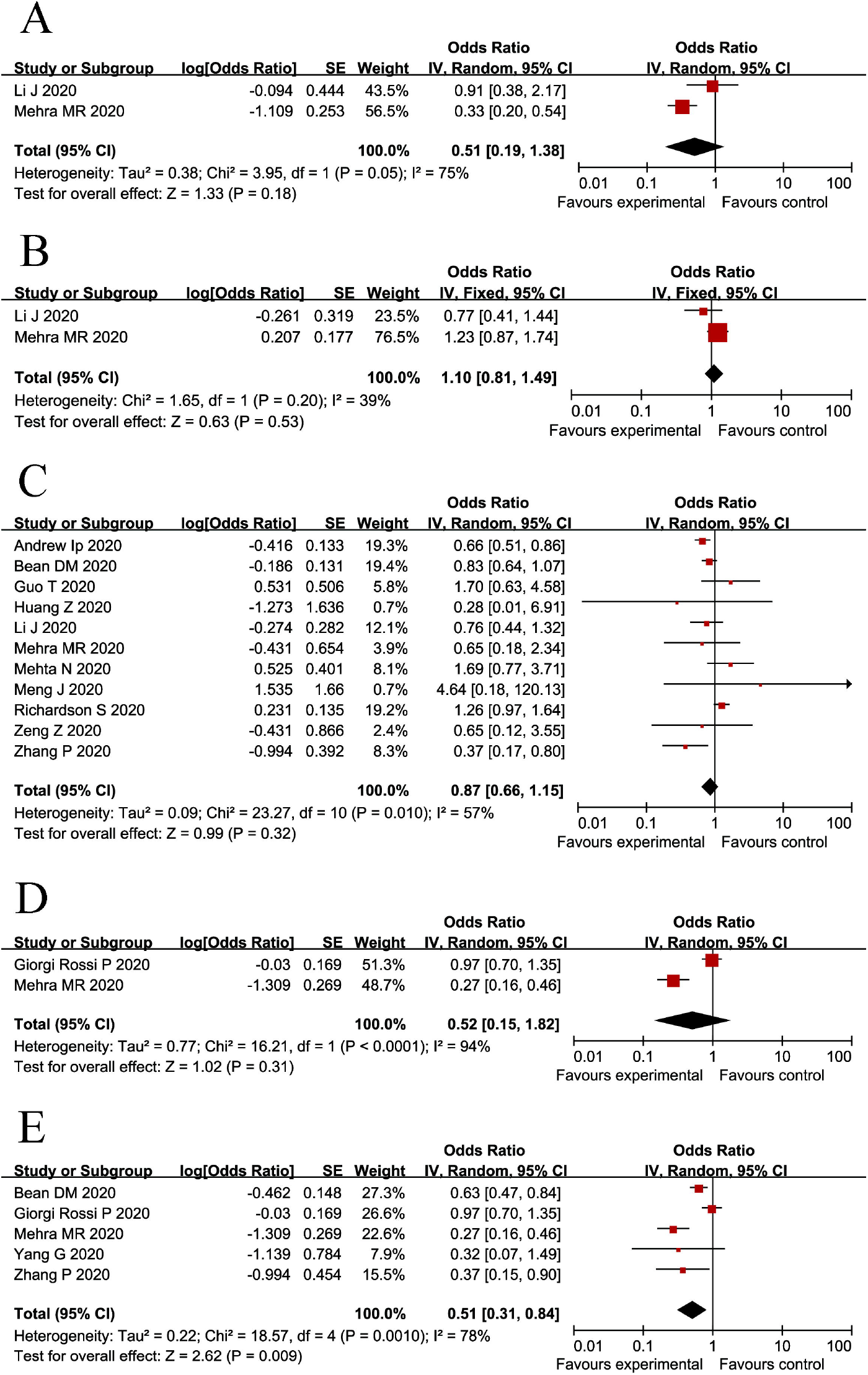
Pooled results of meta-analyses for the association between ACEI/ARB use and risk of death from COVID-19. (A) Pooled crude OR for the comparison between ACEI/ARB use versus non-ACEI/ARB use; (B) Pooled adjusted OR for the comparison between ACEI/ARB use versus non-ACEI/ARB use.

### 4.6 Publication bias and results of sensitivity analysis

There was no significant publication bias among meta-analyses with two or more studies included (All P values were more than 0.05) based on Begg’s and Egger’s tests (Table S4). Sensitivity analysis showed that no study had significant impact on the stability of pooled results from meta-analyses.

## 5. Discussion

The effect of ACEI/ARB use on COVID-19 patients has been a controversial topic since the beginning of this pandemic. Based on prior studies on the effect of ACEI/ARBs on ACE2 expression [7, 38, 39], it was initially hypothesized that increased expression of ACE2 from ACEI/ARB may increase the risk of SARS-COV-2 entrance into respiratory epithelial cells through binding to the structural transmembrane ACE2 receptor. Therefore, ACEI/ARB may theoretically increase the susceptibility of infection and lead to worse outcomes from COVID-19 [11, 40]. However, even though elevated expression of ACE2 caused by ACEI/ARB use has been found in animal studies [7, 38, 39], no evidence has indicated similar effects in human studies. Many studies have been published in the recent months attempting to better understand the effects of ACEI/ARB in COVID-19 patients [8, 10, 19, 29, 30].

Our systematic review and meta-analysis shows that ACEI/ARBs use is not significantly associated with increased infection risk and severity of COVID-19 and is actually associated with a decreased risk of death from COVID-19. However, it does lead to an increased risk of hospital admissions. The increased risk of hospitalization admissions in patients on ACEI/ARB should be interpreted with caution because in the included studies, residual confounding factors such as comorbidities like CVD were not well adjusted for [19, 30]. Patients who are taking ACEI/ARB likely have underlying cardiovascular comorbidities and prior studies demonstrated that patient comorbidities including hypertension and CVD are associated with severe COVID-19 infections, requiring hospitalization, and increased mortality [41, 42]. Therefore, additional large, prospective, randomized studies are needed to better characterize the effect of ACEI/ARB use on rate of hospitalization of COVID-19 patients.

Interestingly, the pooled crude ORs did not show significant association between ACEI/ARB use and the risk of death from COVID-19, but after pooling the adjusted ORs, ACEI/ARB use was associated with a decreased risk of death from COVID-19 in all patients and in those with hypertension. However, the crude OR did not found a significant association of ACEI use and the risk of death from COVID-19. This difference seen bet the crude and adjusted ORs is likely because some important confounders were adjusted for adjusted ORs, which may reflect the true association between ACEI/ARB use and risk of death from COVID-19. Additionally, none of the studies reported the association between ARB use and risk of death from COVID-19; therefore, pooled adjusted OR for ARBs was not calculated.

ACEI/ARBs are commonly used in the treatment of hypertension and CHF to inhibit Angiotensin II and downregulate the Renin-angiotensin-aldosterone system (RAAS) [43]. The decreased risk of death is likely due to continued management of patients’ existing underlying diseases as poorly controlled hypertension and CHF are both associated with worse outcomes in COVID-19 patients [44]. Additionally, the anti-inflammatory and immune-modulatory properties of ACEI/ARBs may also explain their protective effect. Heightened inflammatory response is an important factor leading to adverse outcomes of COVID-19 patients [45]. Previously, it has been shown that Angiotensin II increases expression of inflammatory cytokines via activation of Angiotensin II receptor type 1 (AT1R) [46]. ACEI/ARB decreases the level of Angiotensin II, potentially attenuating the inflammatory response. Additionally, ACEI/ARB reduces T-cell depletion in peripheral blood and inhibits dendritic cell maturation and Th1 and Th17 cell polarization, resulting in a more potent immune system [47]. This hypothesis is supported in the study by Meng et al, in which it was demonstrated that ACEI/ARB use in COVID-19 patients was associated with lower IL-6 level, increased CD3 and CD8 T-cell counts, and decreased peak viral load [25].

This systemic review and meta-analysis comprehensively explored the effect of ACEI/ARB use on infection risk and different clinical outcomes related to COVID-19. Our study is consistent with results from prior studies on this topic, where they also demonstrated that ACEI/ARB use does not increase the severity of COVID-19 infection and decreases mortality [48-50]. Additionally, we also included information not available in previous systemic reviews such as the association between ACEI/ARB use and hospitalization due to COVID-19. This study helps to shed light into the current debate on the use of ACEI/ARBs in patients with COVID-19 and demonstrates that ACEI/ARB use does not lead to increased risk of COVID-19 infection and may even decrease the risk of death from COVID-19. The results from this study supports the joint statement from American Heart Association (AHA), Heart Failure Society of America (HFSA) and American College of Cardiology (ACC) and the statement from the European Society of Cardiology (ESC) Council on Hypertension that patients with COVID-19 taking ACEI/ARBs should continue their treatment [51].

A few limitations present in our study should be noted. First, all the studies included are observational studies, making it difficult to infer accurate causation. Second, no data was provided regarding the dose and exposure duration of ACEI/ARBs; therefore, we could not determinate whether COVID-19 infection is affected by the dose and duration of ACEI/ARBs exposure. Third, beta blockers can also prevent ACE2 activity [52], therefore, the effect of ACEI/ARB use may be underestimated for patients with CVD who also take beta blockers. Finally, given that some important confounders (such as comorbidities) have not been adjusted for in the included studies, the true association between ACEI/ARB use and the risk of hospitalization could not be well inferred.

### Conclusion

In conclusion, ACEI/ARB use does not affect the risk and severity of COVID-19 infection and actually leads to a decreased mortality from COVID-19. The effect of ACEI/ARB use on hospitalization needs to be further assessed with adjusting for potential confounders. The potential protective role of ACEI/ARB supports the recommendation that these agents should not be discontinued for COVID-19 patients if they are already on them.

## Data Availability

Data will be available on request.

## 6. Acknowledgements

Guangbo Qu and Chenyu Sun contributed to the study design, the development of research protocol, and supervision of whole steps of this study. Liqin Shu designed the search strategy and conducted the literature search. Guangbo Qu and Liqin Shu conducted study selection, data extraction and quality assessment of included studies. Chenyu Sun checked the literature search and data extraction. Guangbo Qu performed data analyses and created tables and figures. Guangbo Qu and Chenyu Sun took responsibility for the interpretation of results. Guangbo Qu wrote the first draft of manuscript. Liqin Shu, Evelyn J. Song, Ce Cheng, Dhiran Verghese, John Patrick Uy, Qin Zhou, Hongru Yang, Zhichun Guo, Chenyu Sun contributed to the edition and revision of the draft of manuscript. All authors reviewed and approved final version of the manuscript.

## 7. Funding

This study is financially supported by Hunan Provincial Key Laboratory of Clinical Epidemiology. (2020ZNDXLCL002)

## 8. Disclosure of interest

The authors have no interest to disclosure.

## Notes

### Competing Interest Statement

The authors have declared no competing interest.

### Author Declarations

This manuscript is a collective work of the listed authors, of course, has been read and approved by all the authors. In addition, this paper has not been previously published, not currently under review by another journal and considered for publication elsewhere. All authors have declared no conflict of interest in this study and approved finally version of manuscript.Guangbo Qu and Chenyu Sun contributed to the study design, the development of research protocol, and supervision of whole steps of this study. Liqin Shu designed the search strategy and conducted the literature search. Guangbo Qu and Liqin Shu conducted study selection, data extraction and quality assessment of included studies. Chenyu Sun checked the literature search and data extraction. Guangbo Qu performed data analyses and created tables and figures. Guangbo Qu and Chenyu Sun took responsibility for the interpretation of results. Guangbo Qu wrote the first draft of manuscript. Liqin Shu, Evelyn J. Song, Ce Cheng, Dhiran Verghese, John Patrick Uy, Qin Zhou, Hongru Yang, Zhichun Guo, Mengshi Chen, Chenyu Sun contributed to the edition and revision of the draft of manuscript. All authors reviewed and approved final version of the manuscript

